# Developing a Heart Failure Readmission Model from Inpatient Electronic Medical Record Data

**DOI:** 10.64898/2026.07.18.26358391

**Authors:** Elliot A. Martin, Seungwon Lee, Robin L. Walker, Eda Pitka, Morteza Zangeneh Soroush, Justin A. Ezekowitz, Jonathan G. Howlett, Nowell M. Fine, Jeffrey A. Bakal, Hude Quan, Cathy A. Eastwood

## Abstract

**Importance:** Heart failure readmissions remain common following hospitalization, but accurately identifying which patients will be readmitted after discharge remains challenging. Improved prediction could support targeted transitional care interventions and more efficient allocation of clinical resources.

**Objective:** In this study we attempted to improve readmission prediction after heart failure hospitalization by using variables chosen through a modified Delphi process, and using inpatient Electronic Medical Record (EMR) data, focusing on clinical notes.

**Design:** This prognostic study developed competing risk survival models to predict readmission after heart failure hospitalization. Variables were chosen using a modified Delphi process, and extracted from EMR notes using various natural language processing techniques or from other EMR elements where appropriate. Patients were admitted between 2011 through 2019, and at least one year of follow-up was available for all patients. Models were evaluated using C-statistics, as well as sensitivity, specificity, positive and negative predictive values.

**Setting:** During the study period, all acute-care facilities in Calgary, Alberta used the same EMR system, from which patients were selected.

**Participants:** Patients were 18 years or older, resided in Alberta, and were admitted to a Calgary hospital. All corresponding admissions with a most responsible diagnosis of heart failure were included (n=15,160).

**Main Outcomes and Measures:** The main outcome of interest was readmission within 30 days, though 90– and 365-day time frames were also analyzed. Death was treated as a competing risk and analysed at those time frames as well.

**Results:** Many of the identified variables believed to be important for predicting readmission were not collected reliably enough to be used in readmission models. The final model had a C-statistic of 64.4 when predicting readmission within 30 days, in line with previous studies.

**Conclusions and Relevance:** Efforts to improve heart failure readmission prediction should work with clinical teams to ensure variables believed to be important are collected during hospitalization.

## Introduction

Heart failure (HF) remains a leading cause of hospitalization in North America, including both Canada ^1^ and the United States,^2,3^ and globally.^4^ Thirty-day readmission rates frequently exceed 20%.^1,5^ These readmissions contribute substantially to the economic burden of HF, accounting for a significant share of the ∼$2.8 billion CAN spent annually in Canada^1^, and contributing to the projected $53 billion USD in HF-related costs expected in the United States by 2030.^6^ Readmissions are often considered potentially avoidable, as many patients exhibit recognizable signs of worsening HF prior to hospitalization.^7,8^ Furthermore, HF readmission rates are widely used as indicators of health system performance.^9–11^ As a result, reducing HF admissions has become a major focus of health system quality improvement initiatives.^12,13^

Substantial efforts have been made in recent years to predict HF outcomes after discharge, with a strong focus on readmission ^14,15^ and mortality.^16–18^ The LACE index^19^has been widely implemented due to its simplicity and ease of use^19^; however, it has been validated only with administrative data and consistently demonstrates limited discriminative ability, with a C-statistic below 0.6 for predicting HF readmission^20–23^ Machine learning (ML) approaches have been applied to improve predictive performance, achieving C-statistics generally between 0.6 and 0.8 for readmission, ^14,24–26^ and between 0.7 and 0.9 for mortality.^27–30^ Despite these advances, many studies are limited by methodological issues, including reliance on administrative and clinical trial data not available at the point of care, and the use of models that are difficult to interpret or that are strongly data-driven, which may emphasize variables of limited clinical relevance and be difficult to translate across health systems.

The widespread adoption of electronic medical records (EMR) has generated growing interest in developing HF risk prediction models using routinely collected clinical data. EMRs are designed to support clinical decision-making and have significant potential to accelerate health research,^31^ as they capture granular patient information not available in administrative or clinical trial datasets. Recognizing the potential of EMR data to support predictive modelling, we sought to enhance clinical relevance by conducting a modified Delphi panel to identify clinically relevant variables associated with HF readmission.^32^ This approach allowed us to prioritize variables considered clinically meaningful for HF readmission rather than relying solely on data-driven variable selection. Because the most clinically relevant use of a readmission prediction model is likely at the time of discharge, identifying predictors that are both clinically meaningful and available in real time is particularly important. While prior studies have used EMR data to predict HF outcomes, integrating EMR-derived variables with expert-informed predictors offers an opportunity to improve both predictive performance and clinical applicability. Despite these efforts, accurately identifying patients at high risk of HF readmission remains challenging.

In this study, we developed competing risk models to predict readmission following hospitalization for HF. Our primary objective was to estimate the likelihood of 30-day all-cause readmission post-discharge, treating death as a competing risk. We also evaluated model performance for 90– and 365-day readmission.

## Methods

### Data Sources, Cohort Selection, and Linkage

The HF cohort was defined using the discharge abstract database (DAD), an administrative health dataset that codes for all inpatient encounters. Patients admitted between January 1, 2011, and December 31, 2019, with a most responsible diagnosis of ICD-10-CA I50 were eligible for inclusion. Eligible hospitalizations included patients aged 18 years or older who resided in Alberta and received care in Calgary hospitals operated by Alberta Health Services.

Each eligible hospitalization was treated as a separate index hospitalization for subsequent analyses. Readmission was assessed following discharge from all eligible hospitalizations regardless of discharge disposition.

From this longitudinal dataset, we extracted admission timestamps and constructed episodes of care by combining all admissions to separate acute care institutions within 6 hours of discharge, or within 12 hours when one institution coded a transfer. This process was repeated until no further admissions were added.

We linked the cohort to the Sunrise Clinical Manager™ ^33^ EMR database, which was used across all acute-care facilities in Calgary, Alberta, during the study period. The EMR system contains comprehensive medical data on approximately 1.4 million residents, as well as on individuals hospitalized in Calgary acute-care facilities, regardless of residency. We employed an established linkage mechanism between the DAD and Sunrise Clinical Manager EMR, developed in previous work, ^34^to extract relevant clinical records for the study cohort. The DAD was also used to determine if and when patients were readmitted. The cohort was additionally linked to the Vital Statistics database to determine mortality following the index hospitalization.

This study was approved by the Conjoint Health Research Ethics Board at the University of Calgary (REB20-0684) and received a waiver of consent for this large retrospective cohort. Results are reported using the TRIPOD+AI checklist.^35^

## Extracting Variables of Interest

A previously completed modified Delphi panel involving clinicians, patients, and caregivers identified 61 clinically relevant variables associated with HF readmission.^32^Candidate variables were informed by prior literature and clinical input, and consensus was used to identify those considered important for predicting 30-day all-cause readmission following HF hospitalization. Additional details of the Delphi process are described elsewhere.^32^

These variables were supplemented with additional measures describing the timing of prior ED visits, discharge disposition, and select labs not included by the Delphi panel. Variables available in structured EMR fields, such as lab results and medications, were extracted using standard methods. More complex variables, such as medication adherence, required the use of Natural Language Processing (NLP) techniques. For this study, new NLP algorithms were developed using regular expressions, a rule-based pattern-matching technique. ^36^Where validated algorithms already existed, such as for hypertension status, ^37^ we adopted these approaches. A detailed description of variable extraction methods is provided in Appendix A.

## Competing Risk Modelling & Data Analysis

### Training, Testing, Validation sets, and Missing Values

Variables with more than 5% missingness were excluded, as were highly collinear features (>0.9 Pearson correlation), and only admissions with complete data were included. The dataset was split temporally into three subsets: a training set (2011-2016), a validation set (2017), and a testing set (2018-2019). All numeric columns were scaled to zero mean and unit standard deviation on the training set, and the other subsets were similarly scaled using the training set’s parameters. All models were trained on the training set, with the final model selected based on validation set performance and model complexity. Results are reported exclusively from the test set.

### Modelling, Variable Selection, and Model Selection

Three competing risk models were trained: a Competing Risk Cox model, ^38^ a Fine-Gray model, ^39^ and a Random Survival Forests for Competing Risk model. ^40^ The competing risk Cox and Fine-Gray models were implemented using the R risk Regression package,^41^ while the random survival forests for competing risks were trained using.^40^

Variable selection was performed using recursive feature elimination.^42^ This approach began by fitting a model with all candidate variables, removing the least important variable, and refitting the model with the reduced set of variables. This process was repeated iteratively until the optimal number of features was identified. In the Cox and Fine-Gray models, variable importance was determined by the magnitude of the model coefficients, whereas in the survival forests it was determined by permutation importance.

The optimal number of features was determined by examining plots of the number of features versus the C-statistic and selecting the model with the fewest features before a large drop in the C-statistic. In the Cox and Fine-Gray models, all variables with p-values> 0.05 were removed, and the models were refitted on the training data.

### Statistical Analysis

Descriptive statistics were used to summarize key characteristics of the patient cohort, including demographics, comorbidities, and outcomes. Model discrimination was assessed using C-statistics, sensitivity, specificity, positive predictive value (PPV), and negative predictive value (NPV). Model calibration was evaluated using calibration plots comparing predicted event probabilities with observed event rates.

## Results

The final cohort included 15,160 HF admissions, of which 2,835 (18.7%) were followed by readmission within 30 days and 775 (5.1%) by mortality within 30 days. Summary statistics for the variables included in the final model, as well as for sex, which was not selected, are shown in Table 1. After removing variables with more than 5% missing, as well as highly colinear features, and dropping incomplete records, we were left with 14,374 admissions. The temporal subsets were then: training set (2011-2016; n = 8938; 62%), validation set (2017; n = 1749; 12%), and testing set (2018-2019; n = 3687; 26%).

**Table 1:** Final model variables stratified by readmission, death, and no outcome within 30 days.

|  | Entire Cohort<br>n=15,160 | Readmitted <<br>30 days<br>n=2,835<br>(18.7%) | Died < 30<br>days n=775<br>(5.1%) | No outcome <<br>30 days<br>n=11,828<br>(78.0%) |
| --- | --- | --- | --- | --- |
| Female* | 7,549 (49.8) | 1,412 (49.8) | 383 (49.4) | 5,893 (49.8) |
| Age - median (IQR) | 80.5 (70.4-87.0) | 80.8 (71.6-87.1) | 85.2 (78.2-90.1) | 80.2 (69.8-86.9) |
| Goals of Care (GOC)** |  |  |  |  |
| GOC R1 | 5,106 (33.7) | 839 (29.6) | 81 (10.5) | 4,229 (35.8) |
| GOC R2 | 338 (2.2) | 66 (2.3) | 6 (0.8) | 268 (2.3) |
| GOC R3 | 784 (5.2) | 161 (5.7) | 27 (3.5) | 613 (5.2) |
| GOC M1 | 5,618 (37.1) | 1,242 (43.8) | 278 (35.9) | 4,248 (35.9) |
| GOC M2 | 495 (3.3) | 99 (3.5) | 50 (6.5) | 364 (3.1) |
| GOC C1 | 616 (4.1) | 59 (2.1) | 222 (28.6) | 350 (3.0) |
| GOC C2 | 85 (0.6) | 12 (0.4) | 20 (2.6) | 53 (0.4) |
| GOC Missing | 2,118 (14.0) | 357 (12.6) | 91 (11.7) | 1,703 (14.4) |
| Discharge Disposition |  |  |  |  |
| Acute Care | 770 (5.1) | 121 (4.3) | 37 (4.8) | 622 (5.3) |
| Continuing or Residential Care | 1,210 (8.0) | 186 (6.6) | 180 (23.2) | 880 (7.4) |
| Other (ED/Ambulatory Care/Day<br>Surgery) | 560 (3.7) | 88 (3.1) | 126 (16.3) | 352 (3.0) |
| Home with support | 4,739 (31.3) | 1,090 (38.4) | 251 (32.4) | 3,520 (29.8) |
| Home without support | 7,710 (50.9) | 1,279 (45.1) | 172 (22.2) | 6,359 (53.8) |
| Left against medical advice | 171 (1.1) | 71 (2.5) | 9 (1.2) | 95 (0.8) |
| Previous urgent ER visits in last year - Median (IQR) | 3.0 (1.0-4.0) | 3.0 (2.0-5.0) | 3.0 (2.0-5.0) | 2.0 (1.0-4.0) |
| Previous urgent admissions in last year - Median (IQR) | 1.0 (0.0-2.0) | 1.0 (0.0-2.0) | 1.0 (0.0-2.0) | 1.0 (0.0-2.0) |
| COPD | 5,777 (38.1) | 1,174 (41.4) | 303 (39.1) | 4,412 (37.3) |
| Chronic Kidney Disease | 5,215 (34.4) | 1,205 (42.5) | 335 (43.2) | 3,797 (32.1) |
| Diabetes | 5,426 (35.8) | 1,155 (40.7) | 244 (31.5) | 4,124 (34.9) |
| Anxiety | 3,997 (26.4) | 858 (30.3) | 296 (38.2) | 2,932 (24.8) |
| Depression | 2,470 (16.3) | 482 (17.0) | 140 (18.1) | 1,895 (16.0) |
| Platelets (last measurement) - Median (IQR) | 213.0 (166.0-271.0) | 211.0 (160.0-274.0) | 197.0 (149.0-267.0) | 214.0 (168.0-271.0) |
| Na (last measurement) - median (IQR) | 138.0 (135.0-140.0) | 138.0 (135.0-140.0) | 136.0 (133.0-140.0) | 138.0 (136.0-140.0) |
| Hematocrit (last measurement) - Median (IQR) | 0.4 (0.3-0.4) | 0.3 (0.3-0.4) | 0.3 (0.3-0.4) | 0.4 (0.3-0.4) |

Abbreviations: GOC, Goals of Care; IQR, interquartile range.
All cells are shown as No (%) unless otherwise indicated as median (IQR). All variables except Female were selected by the final model and are significant with $P < 0.05$
\*Female status is included despite not being selected in the final model.
\*\*Definitions of goals of care are provided in https://www.albertahealthservices.ca/frm-103547.pdf.<sup>45</sup> A
corresponding table for 90- and 365-day outcomes is provided Appendix B.

Patients who were readmitted within 30 days had a higher prevalence of chronic kidney disease, diabetes, anxiety, and discharge home with support.

The final model selected was the Competing Risk Cox model with 13 variables, where the categorical variables Goals of Care (GOC) and Discharge Disposition have the results for each category shown in Table 1. Model selection was based on plots of the C-statistic versus the number of features for each model (see Appendix C), and on the performance of the final candidate models on the validation set. As the performance of the Random Survival Forest model was comparable to that of the Competing Risk Cox model, the Cox model was selected for its greater interpretability and familiarity to clinicians. Performance of the final model for predicting readmission on the test set is shown in Table 2. Predictive discrimination was modest across all time horizons, with C-statistics ranging from 64.39 at 30 days to 66.85 at 365 days. At 30 days, the model demonstrated high specificity and low sensitivity.

**Table 2:** Performance Metrics of Final Model at Predicting Readmission on Held Out Test Set.

| Metrics | 30 Days | 90 Days | 180 Days | 365 Days |
| --- | --- | --- | --- | --- |
| C-Stat | 64.39 | 64.71 | 65.6 | 66.85 |
| Sensitivity | 2.56 | 20.06 | 44.73 | 72.77 |
| PPV | 53.12 | 60.61 | 61.59 | 65.51 |
| Specificity | 99.5 | 93.06 | 76.16 | 49.24 |
| NPV | 82.27 | 68.62 | 61.72 | 57.72 |

Figure 1 shows the relationship between predicted readmission risk and predicted mortality risk across the examined time horizons. Each point represents a single admission in the test set.

**Figure 1:**
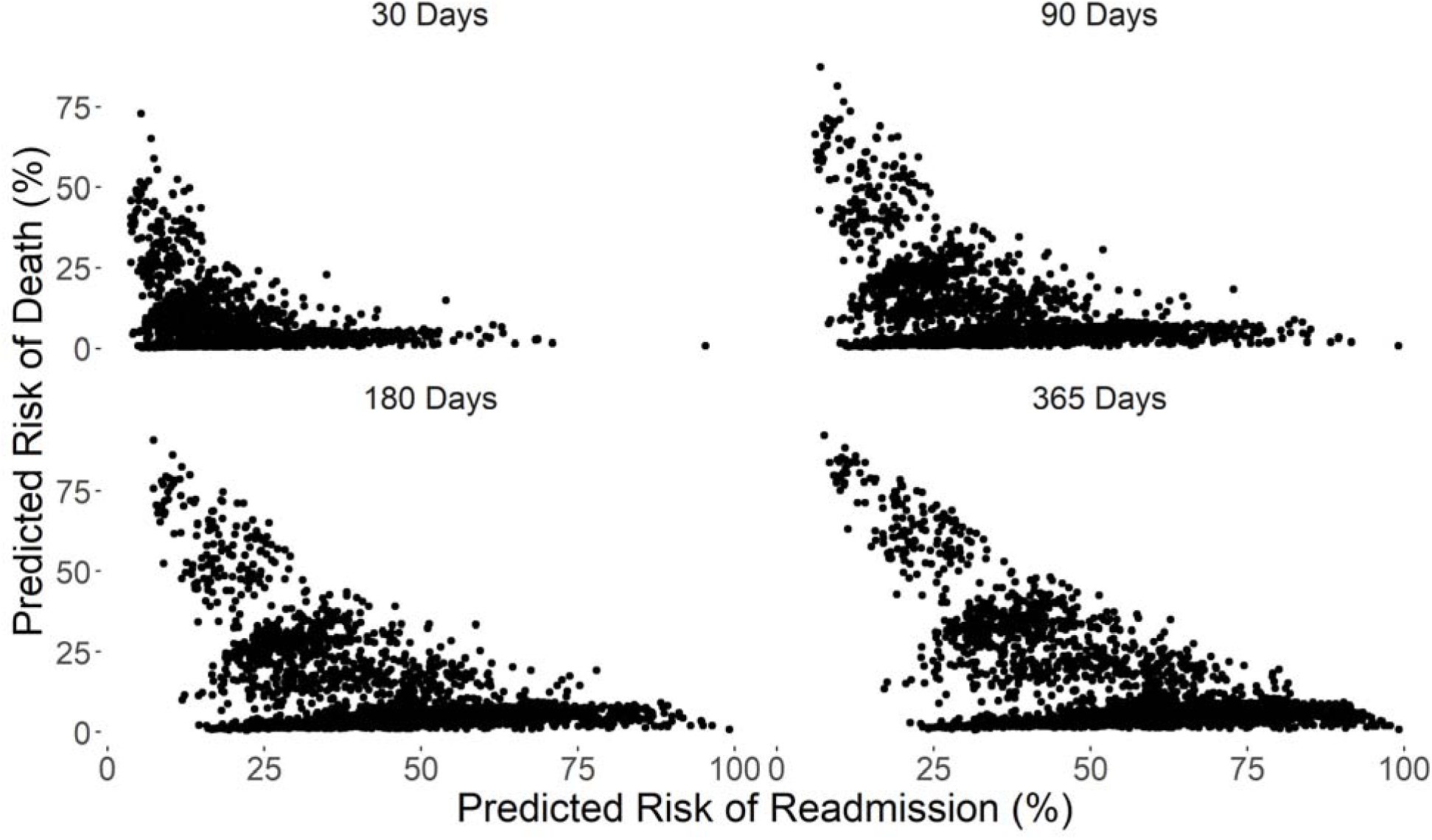
Risk of Readmission vs. Death Each point represents a person in the test set, with predicted risk of readmission on the x-axis and predicted risk of death on the y-axis

Figure 2 shows the calibration of the final model for predicting readmission across the examined time horizons. Calibration plots suggested generally good agreement between predicted and observed risk, although underestimation was observed among lower-risk patients and became more pronounced at longer time intervals.

**Figure 2:**
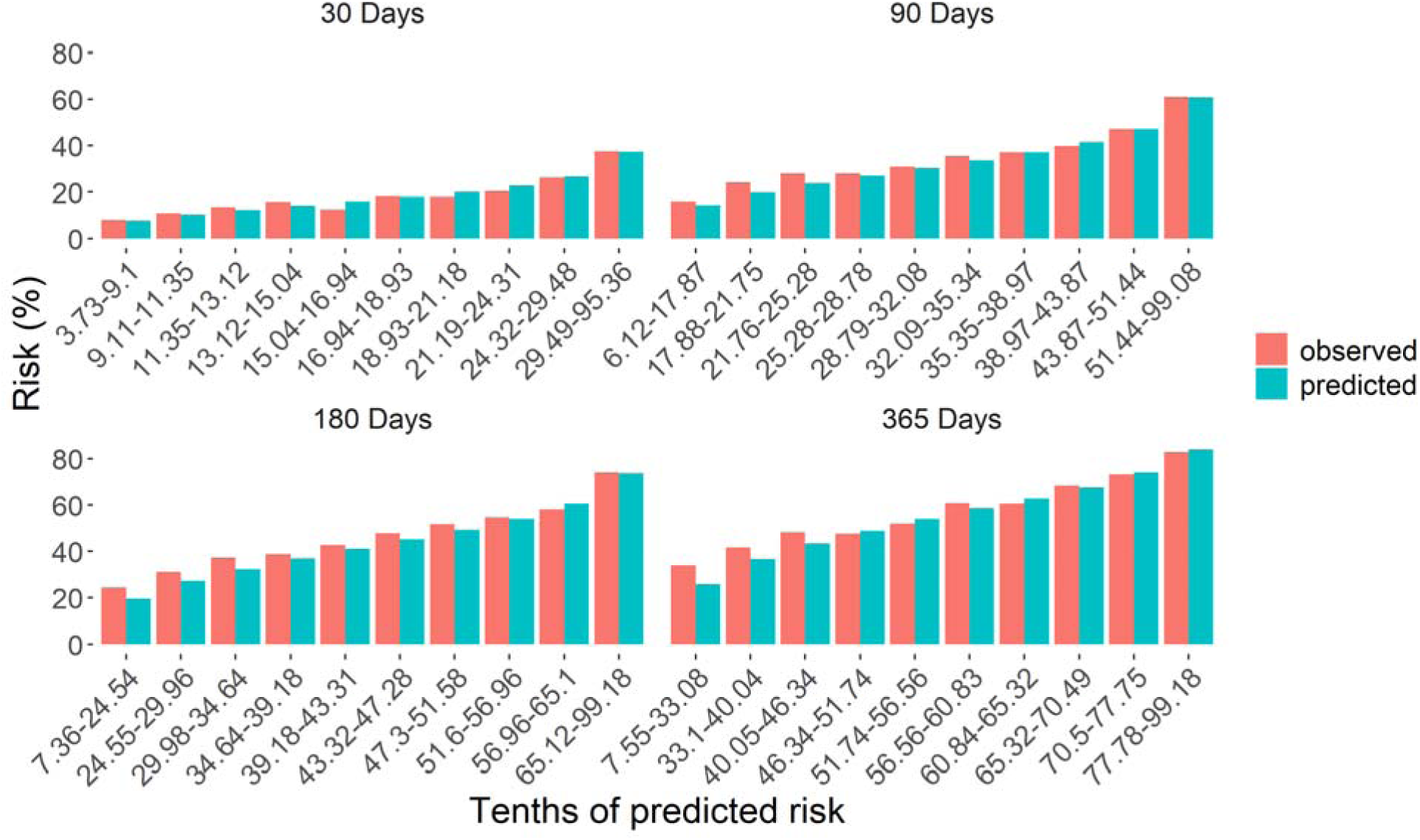
Model Calibration Calibration was assessed by dividing predictions into deciles and plotting the mean predicted risk in each decile (blue bars) alongside the observed event rate (red bars).

## Discussion

Accurately predicting hospital readmission following HF hospitalization remains challenging despite the increasing availability of EMR data and the application of modern modelling techniques. In this study, we developed competing risk models for HF readmission using clinically informed variables derived from linked EMR, hospitalization, and mortality data. Consistent with previous studies evaluating HF readmission prediction models, ^14,15^ the predictive performance of these models was modest, with C-statistics similar to those reported in prior HF readmission prediction models.^14,24–26^ These findings suggest that the difficulty of predicting HF readmission may reflect not only limitations of modelling approaches but also structural limitations in the availability and timing of clinically relevant data within real-world healthcare systems.

An important strength of this work was the use of predictor variables identified through a Delphi panel involving clinicians, patients, and caregivers. This approach allowed us to focus on variables considered clinically meaningful for predicting HF readmission rather than relying solely on data-driven variable selection. However, many of the variables identified as important through this process were excluded from the final models due to missingness in the EMR records. This discrepancy provides insight into some of the practical challenges associated with using routinely collected EMR data for predictive modelling. Notably, prediction of HF mortality has generally demonstrated higher discrimination than prediction of readmission.^14–16,18^ suggesting that factors driving readmission may extend beyond traditional clinical variables.

One key challenge was the limited availability of several clinically relevant variables within the EMR. Several clinically relevant variables identified through the Delphi process were present in fewer than 95% of admissions and were therefore excluded from modelling. For example, biomarkers such as B-type natriuretic peptide (BNP) and blood urea nitrogen (BUN), which are widely used in the diagnosis and management of HF, were missing in 38% and 10% of admissions, respectively, during the study period. Similarly, left ventricular ejection fraction, one of the most important measures used in the clinical classification and management of HF, was rarely documented for the current admission. Although historical ejection fraction measurements could sometimes be identified in prior encounters, these values were often more than one year old and were therefore considered unlikely to reflect the patient’s current clinical status at discharge. Additionally, factors such as the New York Heart Association Functional Classification^43^ and six-minute walk test results^44^ were documented so rarely that they were not extracted. These findings highlight an important gap between variables considered clinically important and those consistently captured in routinely collected data available at discharge.

Several factors identified by the modified Delphi panel that may reduce the likelihood of readmission occur after hospital discharge, such as follow-up with primary care providers or cardiologists. Although these factors are clinically important and may represent potential targets for intervention, they are not available when a model needs to estimate readmission risk for discharge planning and real-time clinical decision-making. In this study, proxy variables were used to approximate some of these processes, such as documentation of a follow-up appointment request. However, these measures cannot capture whether patients ultimately received follow-up care or adhered to treatment recommendations.

More broadly, HF readmissions are influenced by a complex interplay of clinical, behavioral, and health system factors that may not be fully captured within structured EMR data. Factors such as medication adherence, social support, access to outpatient care, and functional status likely play an important role in determining whether patients are readmitted following discharge. Although natural language processing approaches have enabled us to extract some information from free-text clinical documentation, many contextual factors remain difficult to quantify with existing data systems. As a result, prediction models based solely on routinely collected clinical variables may be insufficient to capture the complex determinants of HF readmission.

This study’s findings have important implications for clinicians and health systems attempting to reduce HF readmissions. The most relevant use of a readmission risk model is likely at discharge, when decisions about early follow-up and transitional care planning must be made. In this context, models that depend on post-discharge information may be less useful for real-time clinical decision-making. Although prediction models may help identify patients at elevated risk, our results suggest that discharge-based risk stratification alone may be insufficient to reliably identify those at the highest risk. Efforts to reduce HF readmissions may therefore need to focus more broadly on care transitions, outpatient follow-up, and social and system-level determinants of health rather than relying exclusively on predictive models.

This study has several limitations. First, it was conducted within a single integrated health system in Calgary, Alberta, which may limit the generalizability of the findings to other settings. However, all acute-care hospitals in Calgary used the same EMR during the study period, allowing complete capture of eligible HF hospitalizations within the region. Additionally, the use of clinically informed variables may enhance the relevance of these findings to similar hospital settings. Second, many variables were extracted from free-text EMR documentation using NLP techniques, potentially introducing measurement error. Finally, some clinically important variables were unavailable or inconsistently captured in the EMR, which may have limited model performance.

Consistent with previous studies, our predictive models demonstrated only modest performance despite using clinically informed variables derived from the EMR and a modified Delphi process. These findings suggest that the persistent difficulty of predicting HF readmission may reflect limitations in the availability, completeness, and timing of clinically relevant information within current EMR systems rather than shortcomings of the modelling approaches themselves. Future improvements in prediction may therefore depend less on increasingly sophisticated modelling techniques and more on ensuring that clinically important variables are routinely captured during hospitalization and available at the time of discharge, together with more comprehensive measurement of clinical and social determinants of health.

## Data Sharing Statement

Due to the sensitive nature of the EMR data used in this study we are unable to share it.

## Author Contributions

Drs Martin and Lee had full access to all the data in the study and take responsibility for the integrity of the data and the accuracy of the data analysis.

*Study concept and design*: Eastwood, Quan, Martin, Lee

*Acquisition, analysis, or interpretation of data*: Martin, Lee, Ezekowitz, Howlett, Fine, Bakal, Quan, Eastwood.

*Drafting of the manuscript*: Martin, Lee, Walker, Pitka, Soroush, Eastwood.

*Critical revision of the manuscript for important intellectual content*: Walker, Soroush, Ezekowitz, Howlett, Fine, Bakal, Eastwood

*Statistical analysis*: Martin

*Obtained funding*: Eastwood

*Administrative, technical, or material support*: Pitka, Walker, Soroush.

*Study supervision*: Eastwood, Quan.

## Supporting information

TRIPOD Statement

## Data Availability

Due to the sensitive nature of the electronic medical record data used in this study we are unable to share it.

