## Appendix A for "Developing a Heart Failure Readmission Model from Inpatient Electronic Medical Record Data"

**Appendix A: Delphi Variables and Extraction**

Additional Features not listed in the table but included in the model were, Discharge Disposition, Number of ED visits in the past year, Hematocrit (last measurement taken), and Platelets (last measurement taken).

| **Domain / variable** | **Data source** | **Extraction method** | **Used in final model** | **Reason if excluded** |
| --- | --- | --- | --- | --- |
| **Perihospitalization** | | | | |
| >1 Hospital admission in year prior to hospitalization | Discharge Abstract Database | Determined using the Discharge Abstract Database, where the total count of admissions up to 365 days before the current admission was used as a variable in modeling. | Y |  |
| Multi-disciplinary follow-up post hospitalization | Clinical notes | While it cannot be known if a person will receive multi-disciplinary follow-up at discharge, we can look for evidence that they received a referral for such. This was extracted from clinical notes using a regular expression, knowing the names of the possible clinics they could be referred to, and approximately 37% of patients were documented as having such a referral with this algorithm. | N | Excluded during feature selection |
| Follow-up with primary care provider 1 week post discharge | Clinical notes | As with multi-disciplinary follow-up, it is not known if the patient will follow-up with their primary care provider within 1 week of discharge, so we looked for referrals. This was again done using a regular expression, with approximately 32% of patients having such a referral with this algorithm. | N | Excluded during feature selection |
| Follow-up with cardiologist 1 week post discharge | Clinical notes | As with multi-disciplinary follow-up, it is not known if the patient will follow-up with their primary care provider within 1 week of discharge, so again we looked for referrals. This was again done using a regular expression, however only about 5% of patients had such a referral with this algorithm. | N | Excluded during feature selection |
| Communication of discharge summary to primary care provider | EMR documentation | We explored the EMR documentation to see if we could determine if the discharge summary was communicated to the patient’s primary care provider; however, it did not appear to be feasible. | N | Not feasible |
| Leaving against medical advice during hospitalization | Clinical notes | This was extracted using a regular expression, though only 3% of patients in our cohort were identified as leaving against medical advice using this algorithm. | N | Excluded during feature selection |
| Participation in Advanced Care Planning during hospitalization | EMR system | In our EMR system, advanced care planning was most often captured in Goals of Care (cite). We developed a regular expression to extract the specific goal of care for individual patients on a scale of 1 to 7, with larger numbers corresponding to patients desiring fewer medical interventions. Patients missing a goal of care with our algorithm were labelled as negative 1. | Y |  |
| Receiving nutritional teaching post hospitalization | Clinical notes | It is unknown at discharge if the patient will receive nutritional teaching post hospitalization, so we examined if they received nutritional teaching during their hospitalization. We developed a regular expression to extract if the patient received nutritional teaching during their hospitalization, and approximately 7% of patients were identified as positive for it. | N | Excluded after feature selection as it had a p-value > 0.05 |
| Following a low-sodium diet post hospitalization | Clinical notes | It is unknown at discharge if the patient will follow a low-sodium diet post hospitalization, so we examined if they were following one during their hospitalization. We developed a regular expression to extract if the patient was following a low-sodium diet during their hospitalization, and approximately 8% of patients were identified as positive for it. | N | Excluded during feature selection |
| **Sociodemographics** | | | | |
| Poor Medication Compliance | Clinical notes | This was extractedfrom the clinical notes with a regular expression. Approximately 11% of patients in the cohort were found to have poor medication compliance using the algorithm. | N | Excluded during feature selection |
| Inadequate self-care | Clinical notes | Self-care was found to have many potential dimensions, such as monitoring daily weights, grooming etc., that needed to be captured. Very little documentation was found around these concepts in the clinical notes, so instead we captured “failure to thrive” using a regular expression on the clinical notes. Approximately 4% of patients were documented as failing to thrive with this algorithm. | N | Excluded during feature selection |
| Unemployment (excludes retirement and those on short-term disability or sick leave) | Clinical notes | This was extracted using a regular expression developed in a previous study (cite), though less than 1% of patients in this cohort were labelled as unemployed by this algorithm. | N | Excluded during feature selection |
| Poor/absent fluency in English | Clinical notes | This was extracted using a regular expression developed in a previous study (cite), which was designed to look for language barriers. Approximately 12% of patients in this cohort were labelled as having a language barrier by this algorithm. | N | Excluded after feature selection as it had a p-value > 0.05 |
| Poor or absent prescription drug coverage | Clinical notes | This was extracted using a regular expression, with approximately 8% of patients identified as having drug coverage and less than 1% of patients identified as not having drug coverage. | N | Less than 1% of patients identified as not having drug coverage. As such, this variable was not used in the modeling stage. |
| Chronic disability | Clinical notes | This was determined using a regular expression, where we also tried to use the name of the provincial disability program to help capture patients. This resulted in approximately 9% of patients being labeled as disabled with this algorithm. | N | Excluded during feature selection |
| Unmarried (single, living alone, widowed, or divorced) | Clinical notes | Whether a person was living alone was extracted using a regular expression developed in a previous study (cite), which was designed to look for patients living alone. This was used over marital status, as our previous study found over 12% of patients with a partner were documented as living alone. Approximately 15% of patients in this cohort were labelled as living alone by this algorithm. | N | Excluded during feature selection |
| Age > 75 years | Client table of the EMR | Age was extracted as a structured variable from the client table of the EMR and used as a continuous variable in further analysis (i.e., it was not binarized to over 75 years or not). | Y |  |
| Low income | EMR record | Not applicable | N | As we felt that this would be too missing from the EMR record, we did not develop algorithms for this variable. |
| Smoking (tobacco-like substances) | Clinical notes | This was determined using a regular expression with approximately 18% of patients having a positive diagnosis using this algorithm. | N | Excluded during feature selection |
| Person of colour | EMR system | Not applicable | N | We did not extract this variable as we believed it would be highly under documented in the EMR system. |
| Possessing Indigenous status | Clinical notes | Not applicable | N | We ended up not extracting this variable due to its sensitive nature. |
| **Comorbidities** | | | | |
| Chronic obstructive pulmonary disease | Clinical notes | This was determined using a regular expression, with approximately 38% of patients having a COPD diagnosis using this algorithm. | Y |  |
| Diabetes Type 2 | Clinical notes | This was determined using a regular expression with approximately 36% of patients having a positive diagnosis using this algorithm. | Y |  |
| Atrial fibrillation | Clinical notes | This was determined using a regular expression with approximately 43% of patients having a positive diagnosis using this algorithm. | N | Excluded during feature selection |
| Frailty | EMR system | While searching our EMR system for frailty indications, we found multiple different frailty assessments and scores. | N | We determined that it would be too difficult to reliably label patients as frail at the time, and did not use this variable in the modeling stage. |
| Coronary artery disease (CAD) | Clinical notes | This was determined using a regular expression with approximately 46% of patients having a positive diagnosis using this algorithm. | N | Excluded during feature selection |
| Anemia | Clinical notes | This was determined using a regular expression with approximately 25% of patients having a positive diagnosis using this algorithm. | N | Excluded during feature selection |
| Depression | Clinical notes | This was determined using a regular expression with approximately 16% of patients having a positive diagnosis using this algorithm. | Y |  |
| Anxiety | Clinical notes | This was determined using a regular expression with approximately 26% of patients having a positive diagnosis using this algorithm. | Y |  |
| Substance use disorder (any substance excluding tobacco, marijuana, and vaping) | Clinical notes | This was split into two separate variables, one for alcohol and one for harder drugs such as heroin. Separate regular expressions were developed for each, with approximately 15% of patients identified with alcohol abuse and 2% of patients identified with hard drug abuse with these algorithms. | N | Alcohol use was discarded during feature selection, and hard drugs was discarded as it had a p-value > 0.05 (this could be due to the low proportion of people identified as using hard drugs). |
| Cardiomyopathy | Clinical notes | This was determined using a regular expression with approximately 17% of patients having a positive diagnosis using this algorithm. | N | Excluded during feature selection |
| Atrial Flutter | Clinical notes | This was determined using a regular expression with approximately 8% of patients having a positive diagnosis using this algorithm. | N | Excluded during feature selection |
| Pulmonary embolism/DVT (clotting in the lungs or legs) | Clinical notes | This was determined using a regular expression with approximately 25% of patients having a positive diagnosis using this algorithm. | N | Excluded during feature selection |
| Valve disease | UMLS metathesaurus | The UMLS metathesaurus was used to compile a list of valve diseases and their reported acronyms and synonyms. This was then used to create a regular expression algorithm to identify valve diseases, with approximately 44% of patients having a positive diagnosis using this algorithm | N | Excluded during feature selection |
| Myocardial infarct | Clinical notes | This was determined using a regular expression with approximately 34% of patients having a positive diagnosis using this algorithm. | N | Excluded during feature selection |
| Chronic kidney disease | Clinical notes | This was determined using a regular expression with approximately 34% of patients having a positive diagnosis using this algorithm. | Y |  |
| Cognitive impairment | Clinical notes | This was separated into two variables, dementia and Alzheimer's disease, and separate regular expressions were developed for each. With these algorithms, approximately 12% of patients had a positive dementia diagnosis and 1% of patients had an Alzheimer’s diagnosis. | N | Excluded during feature selection |
| History of hypertension | UMLS CUIs | This variable was extracted using UMLS CUIs (cite) following methods we describe in our previous publication (cite). | N | Excluded during feature selection |
| Congenital heart defect | UMLS metathesaurus | This variable was extracted in a similar fashion to hypertension. Starting with the CUI for Congenital heart defects, all narrower concepts were extracted from the UMLS metathesaurus. We then processed all the clinical notes with cTAKES (cite), and identified anyone with an extracted concept that wasn’t negated and referencing the patient with a congenital heart defect (approximately 4% of patients). | N | Excluded during feature selection |
| Charlson Comorbidity Index >3 | EMR system | Not applicable | N | As this would require us to extract all the Charlson comorbidities from the EMR system, some of which of dubious value e.g., HIV, we did not compute the Charlson comorbidity index. Additionally, we already capture many of the relevant Charlson comorbidities such as COPD, MI, kidney disease etc. |
| **Clinical Features** | | | | |
| Low sodium | EMR system | Sodium measurements were extracted directly from our EMR system. We took the first, last, min, max, and std of the values as potential variables for the models. More than 99% of patients had at least 1 sodium measurement. | Y (the last measurement was chosen) |  |
| High creatinine (Cr) and low GFR | EMR system | Creatinine and GFR measurements were separately extracted directly from our EMR system. We took the first, last, min, max, and std of the values as potential variables of both labs for the models. More than 99% of patients had at least 1 creatinine measurement, and approximately 88% of patients had at least 1 GFR measurement. | N | GFR was excluded as it had more than our 5% missing cutoff, and Cr was excluded during feature selection |
| Six-minute walk test greater than 400 meters | EMR documentation | We explored the EMR documentation for this test but only found 30 patients in our cohort with a result. | N | Due to the very low numbers, we did not go on to extract this variable for use in the models. |
| Tachycardia (high heart rate) | Clinical notes | This was determined using a regular expression with approximately 19% of patients having a positive diagnosis using this algorithm. | N | Excluded during feature selection |
| Peripheral edema (swelling of the extremities) | “Patient Assessment” notes | This was determined using a regular expression that only looked at “Patient Assessment” notes, with approximately 49% of patients having a positive diagnosis using this algorithm. | N | Excluded during feature selection |
| Heart failure with preserved and reduced ejection fraction | echo reports | We developed various algorithms to extract ejection fractions from echo reports but found that most visits did not include an echo. | N | Even allowing for reports up to 6 months before the current visit, we were able to extract an ejection fraction for less than 30% of patients. We decide to not look farther in the past than that, as we believed the result would be too old too be relevant at that point. Therefore, we decided not to utilize ejection fractions in the modeling steps. |
| New York Heart Association functional classification score greater than III | Clinical notes | We explored extracting this variable, but as less than 5% of patients had a score recorded during our analysis, we decided to not extract it for modeling. | N | Not extracted due to lack of documentation found. |
| Low blood pressure | Flowsheets | Both systolic and diastolic blood pressure were extracted from flowsheets using regular expressions. This was found to be highly complete with more than 98% of patients having at least 2 measurements. We took the first, last, min, max, and std of the values as potential variables for the models. | N | Excluded during feature selection |
| Acute kidney failure | Clinical notes | This was determined using a regular expression with approximately 16% of patients having a positive diagnosis using this algorithm. | N | Excluded during feature selection |
| Jugular vein distention | Clinical notes | We explored extracting jugular vein distension but did not extract it for modelling. |  | We found the language around its description to be too varied, with some simply describing it as elevated while others would report the distension in cm relative to different landmarks. |
| **Laboratory** | | | | |
| High brain natriuretic peptide (BNP) or N-terminal BNP | EMR system | In our EMR system the NT-proBNP lab was used. We took the first, last, min, max, and std of the values as potential variables for the models, but they were ultimately dropped in the final analysis as almost 40% of patients did not have this lab result. | N | Excluded before modelling as it was greater than our 5% missing threshold |
| High troponin (HS-cTn) | EMR system | In our EMR system the high sensitivity troponin T lab was used. We took the first, last, min, max, and std of the values as potential variables for the models. We found that approximately 76% of patients had at least one lab result. | N | Excluded before modelling as it was greater than our 5% missing threshold |
| High blood urea nitrogen (BUN) | Clinical notes | For this lab, we took the first, last, min, max, and std of the values as potential variables for the models, but they were ultimately dropped in the final analysis as more than 10% of patients did not have this lab result | N | Excluded before modelling as it was greater than our 5% missing threshold |
| High potassium | EMR system | Potassium measurements were extracted directly from our EMR system. We took the first, last, min, max, and std of the values as potential variables for the models. More than 99% of patients had at least 1 potassium measurement. | N | Excluded during feature selection |
| **Treatments** | | | | |
| Receiving an ARB prescription | Orders for applicable drug names | Prescriptions like this were determined by searching the orders for applicable drug names. Lists of drug names were compiled using ATC codes(cite) to find drug names that belong to the given class of drugs. It was found that approximately 28% of patients had an order for an ARB prescription. |  |  |
| Receiving an ARN-I prescription | Orders for applicable drug names | Prescriptions like this were determined by searching the orders for applicable drug names. Lists of drug names were compiled using ATC codes(cite) to find drug names that belong to the given class of drugs. It was found that approximately 1% of patients had an order for an ARN-I prescription. | N | Excluded after feature selection as it had a p-value > 0.05 |
| Receiving a beta-blocker prescription | Orders for applicable drug names | Prescriptions like this were determined by searching the orders for applicable drug names. Lists of drug names were compiled using ATC codes(cite) to find drug names that belong to the given class of drugs. It was found that approximately 16% of patients had an order for a beta-blocker prescription. | N | Excluded during feature selection |
| Receiving an ACE-I prescription | Orders for applicable drug names | Prescriptions like this were determined by searching the orders for applicable drug names. Lists of drug names were compiled using ATC codes(cite) to find drug names that belong to the given class of drugs. It was found that approximately 47% of patients had an order for an ACE inhibitor prescription. | N | Excluded during feature selection |
| Receiving an SGLT-2 inhibitor prescription | Orders for applicable drug names | Prescriptions like this were determined by searching the orders for applicable drug names. Lists of drug names were compiled using ATC codes(cite) to find drug names that belong to the given class of drugs. It was found that less than 1% of patients had an order for an SGLT prescription. | N | Excluded after feature selection as it had a p-value > 0.05 |
| Receiving an MRA prescription | Orders for applicable drug names | Prescriptions like this were determined by searching the orders for applicable drug names. Lists of drug names were compiled using ATC codes(cite) to find drug names that belong to the given class of drugs. It was found that approximately 29% of patients had an order for an MRA prescription. | N | Excluded during feature selection |
