## Appendix B for "Developing a Heart Failure Readmission Model from Inpatient Electronic Medical Record Data"

### Appendix B: Expanded Table 1

This table includes the demographics and clinical characteristics as listed in Table 1, with additional columns for the outcomes at 90 and 365 days.

|  | Entire Cohort N=15,160 | Readmitted < 90 days n=5,298 (34.9%) | Readmitted < 365 days n=8,854 (58.4%) | Died < 90 days n=1,855 (12.2%) | Died < 365 days n=4,504 (29.7%) |
| --- | --- | --- | --- | --- | --- |
| Female | 7,549 (49.8) | 2,713 (51.2) | 4,508 (50.9) | 941 (50.7) | 2,287 (50.8) |
| Age - median (IQR) | 80.5 (70.4-87.0) | 81.1 (71.8-87.3) | 81.1 (71.5-87.2) | 84.7 (77.5-89.7) | 84.0 (76.3-89.2) |
| Goals of Care (GOC) | |  |  |  |  |
| GOC R1 | 5,106 (33.7) | 1,540 (29.1) | 2,735 (30.9) | 220 (11.9) | 789 (17.5) |
| GOC R2 | 338 (2.2) | 118 (2.2) | 211 (2.4) | 29 (1.6) | 81 (1.8) |
| GOC R3 | 784 (5.2) | 320 (6.0) | 496 (5.6) | 97 (5.2) | 253 (5.6) |
| GOC M1 | 5,618 (37.1) | 2,324 (43.9) | 3,769 (42.6) | 805 (43.4) | 2,116 (47.0) |
| GOC M2 | 495 (3.3) | 181 (3.4) | 292 (3.3) | 124 (6.7) | 245 (5.4) |
| GOC C1 | 616 (4.1) | 104 (2.0) | 161 (1.8) | 340 (18.3) | 461 (10.2) |
| GOC C2 | 85 (0.6) | 21 (0.4) | 32 (0.4) | 37 (2.0) | 53 (1.2) |
| GOC Missing | 2,118 (14.0) | 690 (13.0) | 1,158 (13.1) | 203 (10.9) | 506 (11.2) |
| Discharge Disposition | |  |  |  |  |
| Acute Care | 770 (5.1) | 234 (4.4) | 424 (4.8) | 80 (4.3) | 217 (4.8) |
| Continuing or Residential Care | 1,210 (8.0) | 355 (6.7) | 571 (6.4) | 367 (19.8) | 679 (15.1) |
| Other (ED/Ambulatory Care/Day Surgery) | 560 (3.7) | 168 (3.2) | 269 (3.0) | 186 (10.0) | 317 (7.0) |
| Home with support | 4,739 (31.3) | 1,981 (37.4) | 3,245 (36.7) | 697 (37.6) | 1,739 (38.6) |
| Home without support | 7,710 (50.9) | 2,463 (46.5) | 4,221 (47.7) | 501 (27.0) | 1,494 (33.2) |
| Left against medical advice | 171 (1.1) | 97 (1.8) | 124 (1.4) | 24 (1.3) | 58 (1.3) |
| Previous urgent ER visits in last year - Median (IQR) | 3.0 (1.0-4.0) | 3.0 (2.0-5.0) | 3.0 (2.0-5.0) | 3.0 (2.0-5.0) | 3.0 (2.0-5.0) |
| Previous urgent admissions in last year - Median (IQR) | 1.0 (0.0-2.0) | 1.0 (0.0-2.0) | 1.0 (0.0-2.0) | 1.0 (0.0-2.0) | 1.0 (0.0-2.0) |
| COPD | 5,777 (38.1) | 2,206 (41.6) | 3,674 (41.5) | 776 (41.8) | 1,902 (42.2) |
| Chronic Kidney Disease | 5,215 (34.4) | 2,176 (41.1) | 3,419 (38.6) | 811 (43.7) | 1,932 (42.9) |
| Diabetes | 5,426 (35.8) | 2,077 (39.2) | 3,380 (38.2) | 618 (33.3) | 1,541 (34.2) |
| Anxiety | 3,997 (26.4) | 1,526 (28.8) | 2,432 (27.5) | 651 (35.1) | 1,403 (31.2) |
| Depression | 2,470 (16.3) | 926 (17.5) | 1,531 (17.3) | 362 (19.5) | 818 (18.2) |
| Platelets (last measurement) - Median (IQR) | 213.0 (166.0-271.0) | 211.0 (161.0-271.0) | 211.0 (163.0-270.0) | 201.0 (148.0-265.0) | 203.0 (153.0-264.0) |
| Na (last measurement) - Median (IQR) | 138.0 (135.0-140.0) | 138.0 (135.0-140.0) | 138.0 (135.0-140.0) | 137.0 (134.0-140.0) | 137.0 (134.0-140.0) |
| Hematocrit (last measurement) - Median (IQR) | 0.4 (0.3-0.4) | 0.3 (0.3-0.4) | 0.3 (0.3-0.4) | 0.3 (0.3-0.4) | 0.3 (0.3-0.4) |
