## Appendix C for "Developing a Heart Failure Readmission Model from Inpatient Electronic Medical Record Data"

### Appendix C: Features vs. C-Statistic

The optimal number of features for each model was chosen using recursive feature elimination. Here we show the plots of the number of features each model has vs. the C-statistic of the model. The optimal number of features was chosen as the point where the C-statistic exhibits a pronounced drop. The models were trained on the training set, and results shown are for the validation set.

#### Cox Competing Risk Models

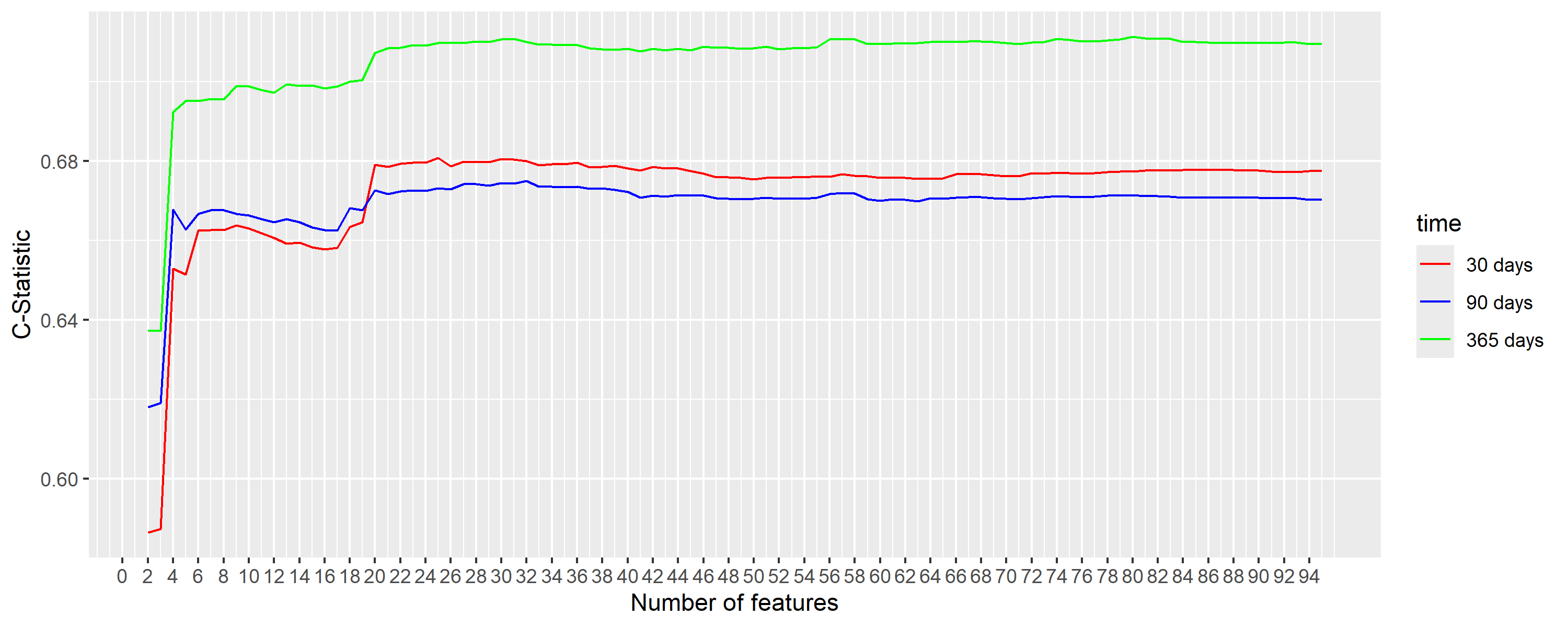

#### Fine Gray Models

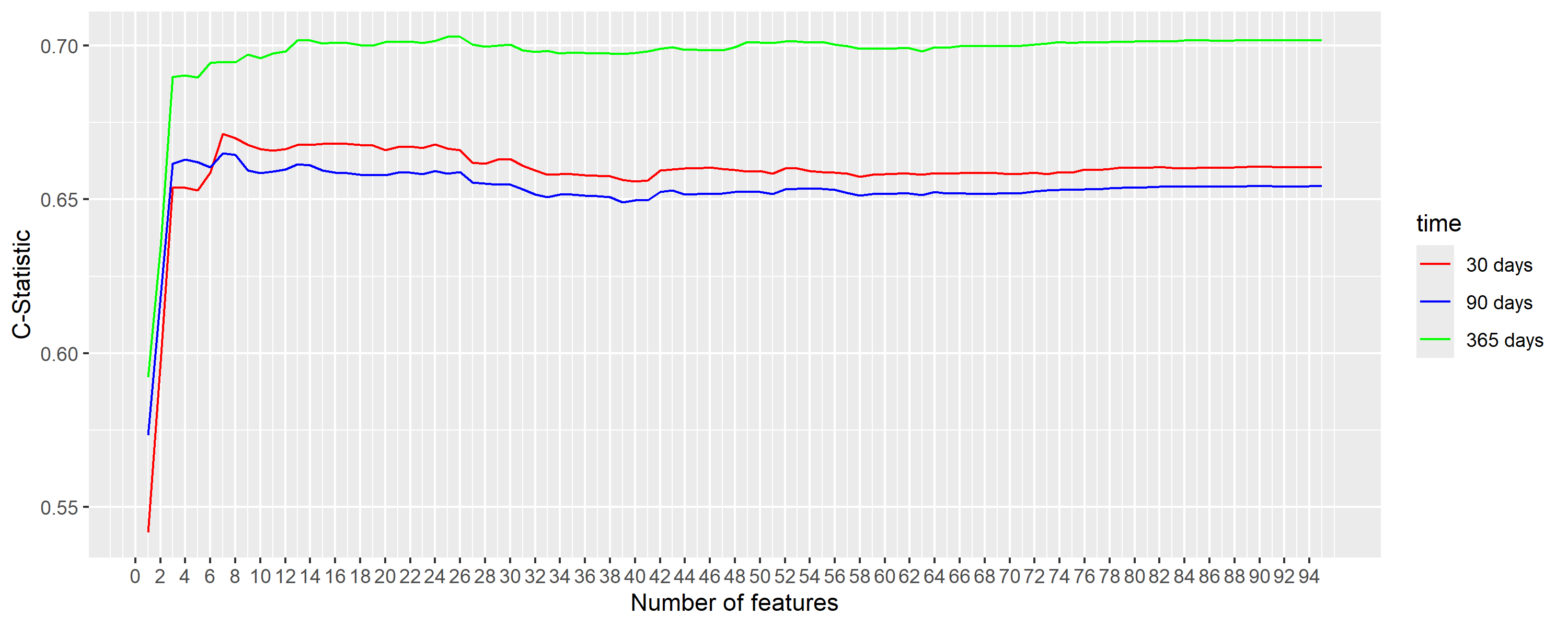

#### Random Survival Forests for Competing Risk Models

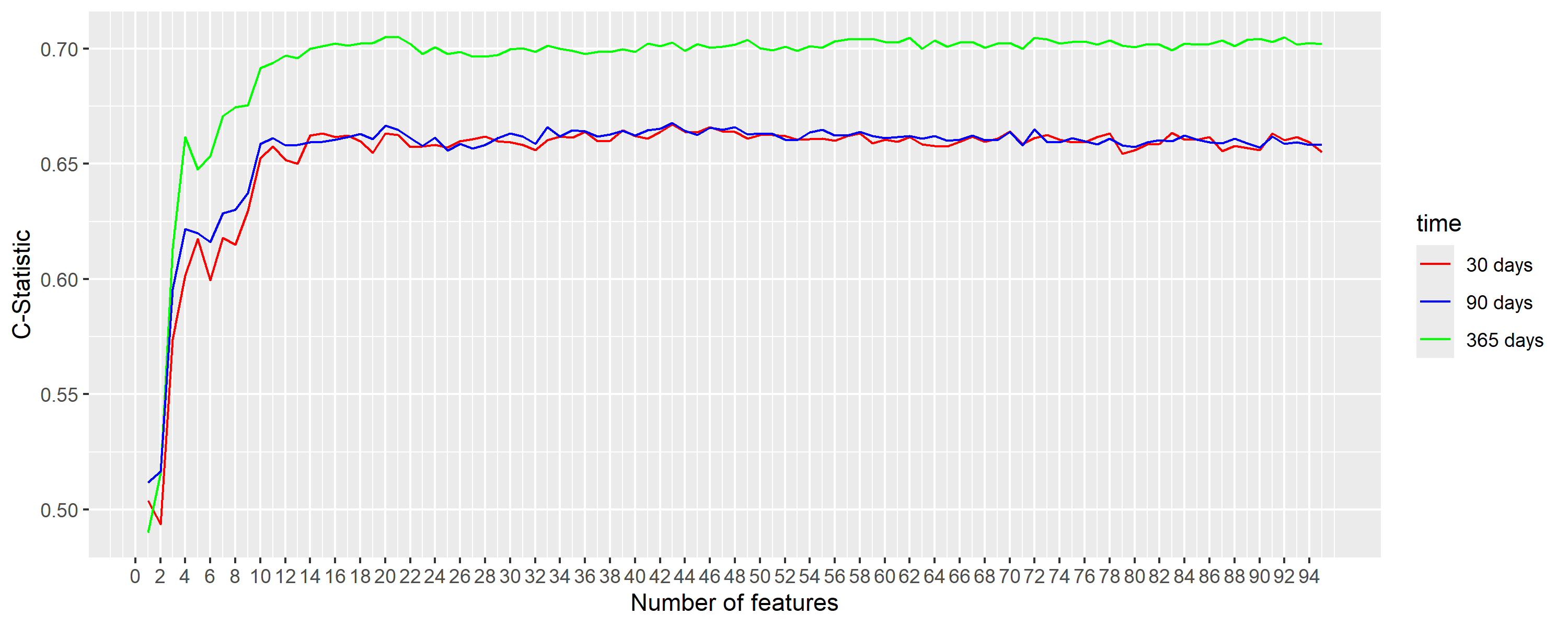
